# Long COVID in hospitalized and non-hospitalized patients in a large cohort in Northwest Spain, a prospective cohort study

**DOI:** 10.1101/2021.08.05.21261634

**Authors:** Alexandre Pérez-González, Alejandro Araújo-Ameijeiras, Alberto Fernández-Villar, Manuel Crespo, Eva Poveda, on behalf of the Cohort COVID-19 of the Galicia Sur Health Research Institute

**Author notes:** **Corresponding autor** Alexandre Pérez González. Galicia Sur Health Research Institute, Group of Virology & Pathogenesis. Internal Medicine Department, Álvaro Cunqueiro Hospital, Vigo. St. Clara Campoamor Nº 341. Zip Code 36312, Vigo, Spain.

## Abstract

**Background:** Survivors to COVID-19 have described long-term symptoms after acute disease. These signs constitute a heterogeneous group named *long COVID* or *persistent COVID*.

**Objective:** The aim of this study is to describe persisting symptoms six months after COVID-19 diagnosis in a prospective cohort in the Northwest Spain

**Design:** This is a prospective cohort study performed in the COVID-19 Cohort of Galicia Sur Health Institute (COHVID-GS).

**Participants:** This cohort includes patients in clinical follow-up in a health area of 569,534 inhabitants after SARS-CoV-2/COVID-19 diagnosis. Clinical and epidemiological characteristics were collected during the follow up.

**Main measures and key results:** A total of 284 patients completed 6 months follow-up, 176 (69.4%) required hospitalization and 29 (10.2%) of them needed critical care. At six months, 119 (48.0%) patients described one or more persisting symptoms. The most prevalent were: extra-thoracic symptoms (39.1%), chest symptoms (27%), dyspnoea (20.6%), and fatigue (16.1%). These symptoms were more common in hospitalized patients (52.3% vs 38.2%) and in women (59.0% vs 40.5%). The multivariate analysis identified Chronic Obstructive Pulmonary Disease (COPD), female gender and tobacco consumption as risk factors for long COVID.

**Conclusions:** Persisting symptoms are common after COVID-19 especially in hospitalized patients compared to outpatients (52.3% vs. 38.2%). Based on these findings, special attention and clinical follow-up after acute SARS-CoV-2 infection should be provided for hospitalized patients with previous lung diseases, tobacco consumption, and females.

## Introduction

COVID-19 pandemic has been a major health challenge around the world, with an unprecedented impact on health systems, society and modern medicine. Acute phase of COVID-19 varies widely, from asymptomatic disease to pneumonia, acute respiratory distress syndrome (ARDS) and multisystem organ failure. Most commonly reported symptoms are fever, cough, dyspnoea, myalgia and anosmia. Several comorbidities have been identified as risk factors for severe disease, such as arterial hypertension, chronic heart disease or diabetes mellitus [1].

In the past few months, many patients reported persisting symptoms after the acute phase of coronavirus disease. These signs are a heterogeneous group, affecting multiple organ systems, including respiratory tract (dyspnoea, chest pain, cough), muscle and joints (arthralgia, myalgia), nervous system (taste or smell alterations, headache, concentration difficulty). There are also many general or unspecified symptoms such as fatigue or hair loss. This group of signs has been named “long COVID” or “persisting COVID”. However, the definition of this syndrome remains unclear and there is not consensus regarding the name and the diagnostic criteria [2]. It is not clear which symptoms could be related to coronavirus and how to define this persistent period after the acute phase [3,4]. The origin of these symptoms remains unknown. However, it is likely to be a combination of a direct damage caused by SARS-CoV-2, previous comorbidities, immunological activation, psychological and emotional factors [5-7]. The aim of this study is to describe persisting symptoms six months after COVID-19 diagnosis in a prospective cohort in the Northwest Spain. The study population includes mild, moderate, serious or critical cases classified as hospitalized or non-hospitalized patients.

## Methods

### Study design

This is a prospective cohort study performed in the COVID-19 Cohort of Galicia Sur Health Institute (COHVID-GS). This is an open cohort associated to a Biobank for sample collection including patients in clinical follow-up at Álvaro Cunqueiro Hospital (Vigo, Spain), a reference hospital for 564,534 inhabitants after SARS-CoV-2/COVID-19 diagnosis (all had a positive PCR for SARS-CoV-2 on nasal swab) (https://www.iisgaliciasur.es/apoyo-a-la-investigacion/cohorte-covid19/). Clinical and epidemiological characteristics were collected at the diagnosis time and during the follow up after 6 months of the diagnosis. The aim of this study is to describe the frequency of long COVID symptoms in hospitalized and non-hospitalized subjects.

### Procedures

All patients included at the COHVID-GS were scheduled for clinical follow-up visits using phone calls and on site visits after 1, 3 and 6 months of the diagnosis. Subjects were classified as hospitalized when they required hospitalization for at least 24 hours. Main hospitalization criteria were pneumonia on chest X-ray and/or respiratory insufficiency. At each visit, patients completed a custom symptom questionnaire (supplementary data). COHVID-GS also includes data about comorbidities, smoke status, weight and height. Pre-existing symptoms that did not change after the diagnosis of COVID-19 were excluded. Symptoms of another suspected origin were also excluded (e.g., iron deficit, thyroid dysfunction). Dyspnoea was classified according to the British Medical Research Council mMRC scale. Brain fog was defined as poor concentration, recent memory disorder or inability to focus.

In this study, we only included patients who survived at least six months after the COVID-19 diagnosis and completed all follow-up visits. We excluded patients who died during the study period, those who refused to complete all follow-up visits and those who were unable to assist to the site (e.g., cognitive impairment, physical disability

The recruitment period started in March 2020 and ended by May 2021 and included consecutive cases. For the purpose of this study, data was censored in January 2021 when the first wave cases reached 6 months since COVID-19 diagnosis.

### Ethics

This project was approved by the Ethical Committee of Pontevedra-Vigo-Ourense (reference 2021/184). All the patients included in the study belong to the COHVID-GS, which signed an informed consent form at the time of inclusion in the cohort.

### Statistical analysis

For quantitative variables, median and interquartile were calculated. Categorical variables were reported as frequencies and percentages. Subjects were classified in two groups, based on their need for hospitalization due to Covid-19. Symptoms, clinical and epidemiological data were compared in both groups using chi-square test, Fisher’s exact test or U Man-Whitney test as appropriate. P-value less than 0.05 was considered significant. Multivariable logistic regression analysis with backward stepwise elimination was used to identify risk factors for long COVID. Those variables with a p value lesser than 0,1 were considered as independent variables. Statistical analyses were performed with IBM Statistical Product and Service Solutions (SPSS) program, version 22.

## Results

### Baseline characteristics of the study population

A total of 630 patients were included in the COHVID-GS. However, 81 of them died during the hospitalization period and only 284 subjects who survived completed the six months period of follow-up. Of them, 172 required hospitalization and 76 were never hospitalized. The baseline characteristics of the study population are shown in **Table 1**.

**Table 1.** Baseline characteristics of study population

|  | **Total** | **Hospitalized** | **Non hospitalized** | **p-value** |
| --- | --- | --- | --- | --- |
| **Patients in follow-up, n (%)** | 248 (100%) | 172 (69.4%) | 76 (30.6%) |  |
| **Demographics** |  |  |  |  |
| **Age, in years, median (IQR)** | 57 (46—68) | 62 (51—71) | 47 (34 – 54) | $p<0.001$ |
| **Sex, male, n (%)** | 148 (59.7%) | 103 (59.9%) | 45 (62.5%) | $p=1.000$ |
| **Spanish, n (%)** | 234 (94.4%) | 159 (92.4%) | 75 (98.7%) | $p=1.000$ |
| **Latin American, n (%)** | 11 (4.4%) | 10 (5.8%) | 1 (1.3%) | $p=0.070$ |
| **European (not Spanish), n (%)** | 2 (3.8%) | 2 (1.2%) | 0 | $p=0.267$ |
| **Oceania, n (%)** | 1 (0.4%) | 1 (0.6%) | 0 | $p=1.000$ |
| **Severity** |  |  |  |  |
| **Admission to critical care, n (%)** | 29 (11.7%) | 29 (16.8%) | N/A | N/A |
| **Invasive mechanical ventilation required, n (%)** | 23 (9.3%) | 23 (13.4%) | N/A | N/A |
| **Comorbidities** |  |  |  |  |
| **Obesity, n (%)** | 75 (30.2%) | 62 (36.0%) | 13 (17.1%) | $p<0.001$ |
| **Arterial hypertension, n (%)** | 65 (26.2%) | 58 (33.7%) | 7 (9.2%) | $p<0.001$ |
| **Diabetes mellitus, n (%)** | 26 (10.5%) | 25 (14.5%) | 1 (1.3%) | $p=0.001$ |
| **Asthma, n (%)** | 21 (8.5%) | 7 (4.1%) | 14 (18.4%) | $p=0.807$ |
| **Chronic ischaemic heart disease, n (%)** | 21 (8.5%) | 21 (12.2%) | 0 | $p<0.001$ |
| **Chronic obstructive pulmonary disease, n (%)** | 15 (6.0%) | 15 (8.7%) | 0 | $p=0.007$ |
| **Active cancer, n (%)** | 5 (2.0%) | 4 (2.3%) | 1 (1.3%) | $p=1.000$ |
| **HIV infection, n (%)** | 3 (1.2%) | 2 (1.2%) | 1 (1.3%) | $p=1.000$ |
| **Chronic kidney disease, n (%)** | 3 (1.2%) | 3 (1.7%) | 0 | $p=0.555$ |
| **Liver cirrhosis, n (%)** | 2 (0.8%) | 2 (1.2%) | 0 | $p=1.000$ |
| **Tobacco use** |  |  |  |  |
| **Active smoker, n (%)** | 18 (7.3%) | 8 (4.6%) | 10 (13.9%) | $p=0.031$ |
| **Former smoker, n (%)** | 105 (42.3%) | 75 (43.6%) | 35 (48.6%) | $p=0.575$ |
| **Never smoker, n (%)** | 120 (69.8%) | 85 (49.4%) | 35 (20.3%) | $p=0.582$ |

Overall, the median age was 57 years, however, non-hospitalized patients were significantly younger than those who were hospitalized (62 vs. 47 years, respectively). Moreover, male patients were more frequent in both groups (148 males, 59.7%).

Most common comorbidities were obesity (n=75, 30.2%), followed by arterial hypertension (n=65, 26.2%) and diabetes mellitus (n=26, 10.1%). Moreover, all comorbidities were more usual in the hospitalized group, except for asthma (4.1% vs. 18.4% respectively). In the hospitalized group, 29 patients (16.8%) required critical care management, including 23 who required invasive mechanical ventilation.

Six months after the COVID-19 diagnosis, 119 patients (48.9%) reported at least one symptom (**Table 2**). Chest symptoms were more frequent (n=67, 27%), of which dyspnoea (n=51, 20.6%), chest pain (n=15, 6.0%) and cough (n=11, 4.4%) were the most commonly reported.

**Table 2.**
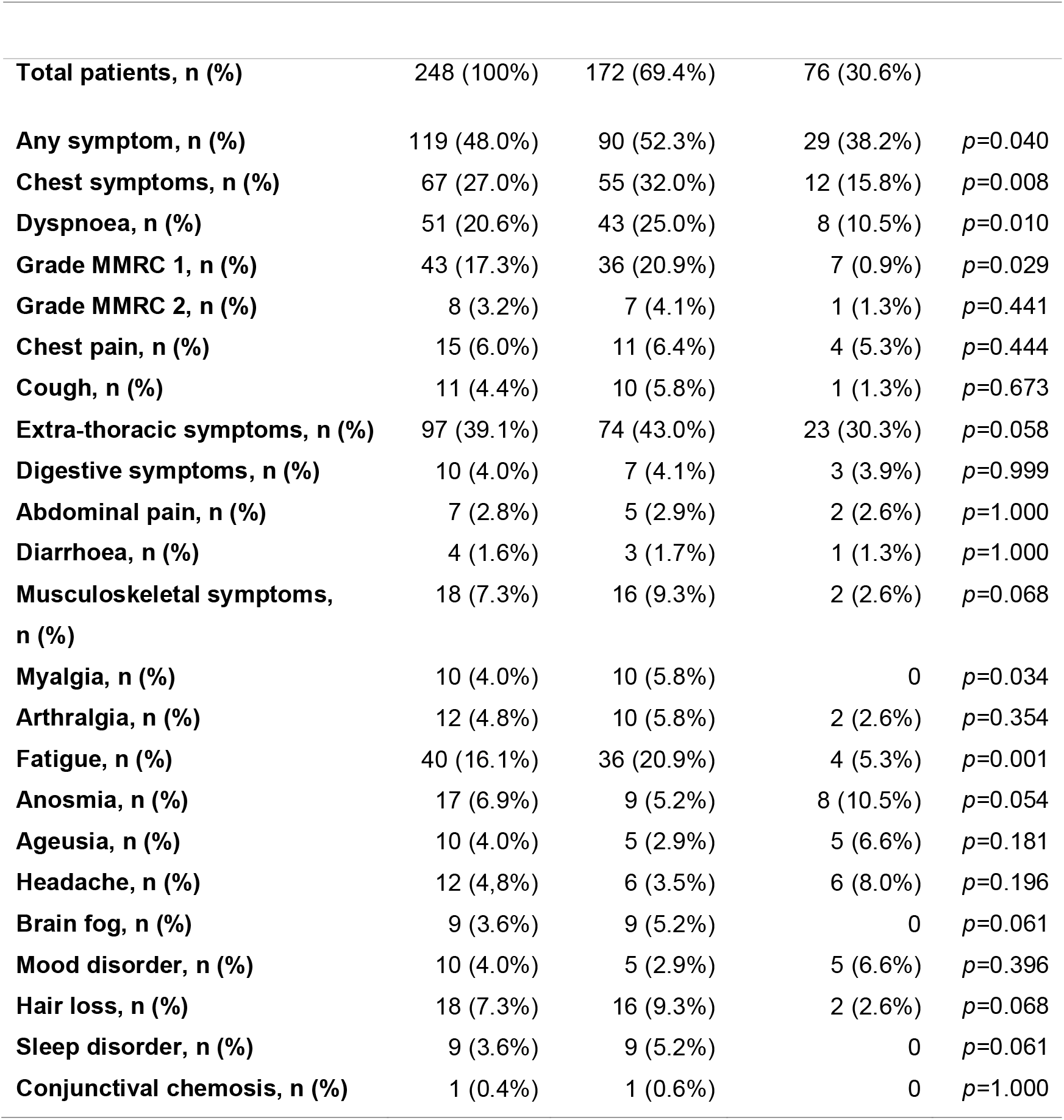
Prevalence of 6 months persisting symptoms

Extra-thoracic symptoms were described in 97 patients (39.1%) and the most frequently reported were fatigue (n=40, 16.1%), musculo-skeletal symptoms (n=18, 7.3%) and hair loss (n=18, 7.3%).

Overall, smell disorders were more frequent than taste disorders (6.9% vs. 4.0%, respectively). Neurological symptoms six months after the COVID-19 diagnosis, excluding smell and taste disruptions, were uncommon (less than 5.0%). Headache, mood disorder and sleep disturbance were the most common neurological symptoms. Nine patients (3.2%) reported symptoms that met *brain fog* criteria. Digestive symptoms were also uncommon (n=10, 4.0%).

### Persisting symptoms in hospitalized and non-hospitalized patients

Persisting symptoms were more prevalent in the hospitalized group (52.3% vs. 38.2%, p=0.040). Chest symptoms in this group were more frequently reported compared with the non-hospitalized group (32.0% vs. 15.8%, p=0.008). However, extra-thoracic symptoms did not differ between both groups (43.0% vs. 30.3%, p=0.058). The most common chest symptom was dyspnoea, which was more frequently reported in hospitalized patients (25.0% vs. 10.5%, p=0.010). Most of the subjects reported a mMRC (modified Medical Research Council) grade 1 dyspnoea (n=43, 17.3%). On the other hand, chest pain (6.4% vs. 5.3%) and persisting cough (5.8% vs. 1.3%) did not vary between both groups.

Fatigue (20.9% vs. 5.3%, p=0.001) and myalgia (5.8% vs. 0%, p=0.034) were more frequently reported in the hospitalized group than in the non-hospitalized group. Also, hair loss (9.3% vs. 2.6%) and sleep disorders (5.2% vs. 0%) were more common in hospitalized patients, although this difference was not statistically significant. Arthralgia (5.8% vs. 2.6%), ageusia (2.9% vs. 6.6%) and digestive symptoms (4.1% vs. 3.9%) did not differ between both groups. Anosmia was more prevalent in non-hospitalized subjects (5.2% vs. 10.5%, p=0.054), but did not meet statistical significance.

### Persisting symptoms in male and female patients

Overall, persisting symptoms were more common in females than males (59.0% vs. 40.5%, p=0.004) (**Table 3**). Dyspnoea (29.0 vs. 14.9%, p=0.007), headache (10.0% vs. 4.8%, p=0.004), fatigue (22.0% vs. 12.2%, p=0.039) and hair loss (15.0% vs. 7.3%, p<0.001) were all more prevalent in females while anosmia (6.0% vs. 7.4%), taste disturbance (4.0% in both genders) and musculoskeletal symptoms (6.8% vs. 8.0%) did not differ between females and males.

**Table 3.** Prevalence of persisting symptoms at 6 months based on gender

|  | Female | Male | p-value |
| --- | --- | --- | --- |
| **Any symptom, n (%)** | 59 (59.0%) | 60 (40,5%) | $p=0.004$ |
| **Chest symptoms, n (%)** | 35 (35.0%) | 32 (21,6%) | $p=0.020$ |
| **Dyspnoea, n (%)** | 29 (29.0%) | 22 (14.9%) | $p=0.007$ |
| **Chest pain, n (%)** | 7 (7.0%) | 8 (5.4%) | $p=0.600$ |
| **Cough, n (%)** | 7 (7.0%) | 4 (2.7%) | $p=0.125$ |
| **Extra-thoracic symptoms, n (%)** | 50 (50.0%) | 47 (31.8%) | $p=0.004$ |
| **Neurological symptoms, n (%)** | 26 (26%) | 21 (14.2%) | $p=0.022$ |
| **Anosmia, n (%)** | 6 (6.0%) | 11 (7.4%) | $p=0.800$ |
| **Agueusia, n (%)** | 4 (4.0%) | 10 (4%) | $p=1.000$ |
| **Headache, n (%)** | 10 (10%) | 12 (4.8%) | $p=0.004$ |
| **Brain fog, n (%)** | 5 (5%) | 4 (2.7%) | $p=0.491$ |
| **Diarrhea, n (%)** | 4 (4.0%) | 0 | $p=0.025$ |
| **Myalgia, n (%)** | 5 (5.0%) | 5 (3.4%) | $p=0.530$ |
| **Arthralgia, n (%)** | 4 (4.0%) | 8 (5.4%) | $p=0.767$ |
| **Fatigue, n (%)** | 22 (22%) | 18 (12.2%) | $p=0.039$ |
| **Hair loss, n (%)** | 15 (15%) | 18 (7.3%) | $p<0.001$ |

### Predictors of COVID-19 persisting symptoms

We performed a multivariate analysis in order to determine the risk factors for persisting COVID-19 symptoms. We identified Chronic Pulmonary Obstructive Disease (COPD) (OR 5.009; CI 95%, 1.321 – 18.997, p=0.018), female gender (OR 2.700, 95% CI 1.543 – 4.724, p<0.001) and tobacco consumption as independent risk factors for remaining symptoms (OR 1.736; CI 95%, 1.002 – 3.007, p=0.049) (**Table 4**).

**Table 4.** Risk factors for long COVID.

|  | **Odds Ratio** | **Confidence interval (95%)** | **p-value** |
| --- | --- | --- | --- |
| **COPD** | 5.009 | 1.321 – 18.997 | $p=0.018$ |
| **Sex, female** | 2.700 | 1.543 – 4.724 | $p<0.001$ |
| **Hospitalization** | 1.711 | 0.951 – 3.078 | $p=0.073$ |
| **Tobacco consumption** | 1.736 | 1.002 – 3.007 | $p=0.049$ |

*COPD: chronic obstructive pulmonary disease*

COPD was the only comorbidity associated with a higher prevalence of persisting symptoms. Fourteen patients (5.1%) were affected by COPD before COVID-19 diagnosis and 12 of them (80.0%) had at least one persisting symptom. Nine subjects (75.0%) reported new or worsened dyspnoea compared with their perception prior COVID-19 diagnosis. Dyspnoea (60.0% vs. 18.0%, p=0.001) and chest pain (26.7% vs. 4.7%, p=0.008) were most commonly reported in COPD group. On the other hand, cough (6.7% vs. 4.3%), fatigue (20.0% vs. 15.9%), myalgia (13.3% vs. 3.4%), arthralgia (6.7% vs. 4.7%) did not differ between COPD and non-COPD subjects.

## Discussion

We characterized long COVID in a prospective cohort of patients with mild and severe COVID-19 with a systematic clinical follow-up. The prevalence of any symptom at six months was elevated (48%), with a different distribution between hospitalized and non-hospitalized patients. Persisting symptoms did not differ between those patients who needed critical care and those who did not.

Long COVID has been recognized as a public-health problem and a matter of concern for COVID-19 surveys. Several studies have reported information in series of patients in clinical follow-up after the acute infection, but many questions remain unsolved. One of them is if long COVID might be gender-dependent. In our study, females showed a higher prevalence of persisting symptoms in hospitalized and in non-hospitalized groups. Dyspnoea, headache, fatigue and hair loss were all more common in females than males. A prior study performed by Chaoling-Huang et al., also described a higher frequency of long COVID symptoms in females than males [8]. To the best of our knowledge, there are only few studies evaluating long COVID symptoms between males and females, both in hospitalized and non-hospitalized setting [10].

In the recent months, more than 200 persistent symptoms have been described after the acute phase of COVID-19. Most of them are related to the respiratory tract (dyspnoea, cough, chest pain), the main target of SARS-CoV-2. However, many patients report other type of symptoms such as fatigue, cognitive impairment or smell disorders [11].

Chest symptoms were the most common reported affection, with greater prevalence in the hospitalized group. This difference may be related to pneumonia, respiratory distress and lung damage, which were more frequent in hospitalized subjects. The prevalence of chest symptoms in this group raised to 52%, slightly higher than observed in previous reports [9]. However, even in the absence of pneumonia, 15% of non-hospitalized patients reported persisting chest symptoms. Recently, Blomberg et al., described long COVID symptoms in Norwegian non-hospitalized and hospitalized population with a slightly higher prevalence than in our study (52.0% vs. 38.2% respectively) [10].

Fatigue was also frequent, reaching up to 20.9% in the hospitalized group. Although this percentage is relevant, it was lower than others reported before [12,13], around 65 – 70%. However, these studies had a shorter follow-up period (90 to 100 days after hospitalization). The reason of fatigue remains unknown, but it is probably a result of a combination of factors, including direct nervous and muscle damage due to SARS-CoV-2 replication [5,6], immune activation and immune dysregulation, or emotional factors [7].

On the other hand, myalgia and arthralgia were less frequent than in previous studies. Sykes et al., described a prevalence of 50% of these symptoms in hospitalized patients at 100 days after discharge [14].

Overall, persisting anosmia was reported in 6.8% of subjects and was more common in the non-hospitalized group (10.5% vs. 5.2%, respectively). This prevalence is lower than published in previous studies [15, 16]. In addition, prior reports have suggested that anosmia may be associated with a mild to moderate COVID-19 and it could be more common in females than males [15]. However, the low prevalence of anosmia observed in our study cannot confirm this hypothesis.

At six months, 9 subjects reported unspecified neurological symptoms such as poor concentration, loss of recent memory or inability to focus. These unspecified symptoms are named *brain fog* by some authors, but its origin or relationship with coronavirus disease remains unknown [5, 17]. *Brain fog* syndrome after COVID-19 has been reported previously [18, 19], although there is not a validated diagnostic criteria established. The studies focused on neurological *sequelae* of COVID-19 did not use a unanimous criterion and therefore, the conclusions are very heterogeneous. The lack of knowledge about this type of symptoms makes more difficult to study the real prevalence or to establish a relationship with coronavirus disease. Another neurological symptoms like sleep or mood disorders have been reported with a similar prevalence than in our study [20]. However, psychological conditions, stress and isolation may be associated with these symptoms, alongside with coronavirus infection. The higher prevalence on the hospitalized group may be due to the severity of illness, admission to critical care or treatments (e.g., corticosteroids, immunosuppressants).

We identified COPD, female gender, tobacco consumption and the need for hospitalization as predictors for long COVID. Female gender has been reported before as a risk factor for hospitalized subjects but there is a lack of information about other predictors of long COVID.

Our findings suggest that some of the persisting symptoms could be related to a previous lung comorbidity and hospitalization. SARS-CoV-2 damage on respiratory tract could be more intense in patients with previous lung disease, increasing the risk of long-term symptoms. In addition, pneumonia and ARDS may also contribute to develop persisting symptoms.

This study has few limitations. Firstly, some of the symptoms reported are subjective and based on the patient’s testimony (e.g. fatigue, headache, dyspnoea). Additionally, the lack of validated scales to measure most of the symptoms makes difficult to compare data between subjects or studies. Secondly, most of the symptoms may be affected by personal, psychological or environmental factors. We needed to use a non-validated custom questionnaire, as there is not a specific post-COVID-19 document established. Many symptoms could be higher reported in one group than another for unrelated reasons with coronavirus disease. This is the case for hair loss that may be more noticed in women and therefore more reported, leading to a possible increase of the prevalence in female subjects. In addition, previous comorbidities and age could increase the persisting symptoms in the hospitalized group. For COPD patients, we did not record symptoms that started before COVID-19 diagnosis. Therefore, it is possible that COPD patients reported more frequently chest or respiratory symptoms.

## Conclusions

In summary, a high prevalence of long COVID (48%) was observed in patients after six months of acute SARS-CoV-2 infection. Overall, persistent symptoms were more common in hospitalized patients compared to outpatients (52.3% vs 38.2%). These symptoms were more common in females than males (59.0% vs. 40.5%). COPD, female gender, and tobacco consumption were identified as long COVID predictors. These findings suggest that in those patients that required hospitalization after SARS-CoV-2 infection a close clinical follow-up might be recommended. Special attention should be provided for patients with previous lung diseases, tobacco consumption and females.

## Supporting information

Questionnaire

## Data Availability

The data that support this study cannot be publicly shared due to ethical or privacy reasons and may be shared upon reasonable request to the corresponding author if appropriate.

## Acknowledgments

We would like to thank all the members of COHVID-GS and IISGS Biobank, patients and nursing staff and the Methodology and Statistic Unit.

## Declaration of interests

The authors declare no competing interests.

## Members of COHVID-GS (Galicia Sur Health Research Institute)

Alejandro Araujo, Jorge Julio Cabrera, Víctor del Campo, Manuel Crespo, Alberto Fernández, Beatriz Gil de Araujo, Carlos Gómez, Virginia Leiro, María Rebeca Longueira, Ana López-Domínguez, José Ramón Lorenzo, María Marcos, Alexandre Pérez, María Teresa Pérez, Lucia Patiño, Sonia Pérez, Silvia Pérez-Fernández, Eva Poveda, Cristina Ramos, Benito Regueiro, Cristina Retresas, Tania Rivera, Olga Souto, Isabel Taboada, Susana Teijeira, María Torres, Vanesa Val, Irene Viéitez

## Funding

COVID-19 Cohort Galicia Sur Health Research Institute (COHVID-GS) is funded by the Instituto de Salud Carlos III through the FONDO COVID19 (COV20/00698). Alexandre Pérez is hired under a Rio Hortega contract (CM20/00243).

## Data availability

The datasets during and/or analyzed during the current study available from the **COHVID-GS** on reasonable request (https://www.iisgaliciasur.es/apoyo-a-la-investigacion/cohorte-covid19/).

## Notes

### Competing Interest Statement

The authors have declared no competing interest.

### Funding Statement

This research did not receive any specific grant from funding agencies in the public, commercial, or not-for-profit sectors. Alexandre Perez, principal investigator, is hired under a Rio Hortega contract financed by Instituto de Investigacion Carlos III (ISCIII) with reference number CM20/00243.

## References

[1] Berenguer J, Ryan P, Rodríguez-Baño J, et al. Characteristics and predictors of death among 4035 consecutively hospitalized patients with COVID-19 in Spain. Clin Microbiol Infect. 2020;26(11):1525–1536. doi:10.1016/j.cmi.2020.07.024

[2] Al-Jahdhami I, Al-Naamani K, Al-Mawali A. The Post-acute COVID-19 Syndrome (Long COVID). Oman Med J. 2021;36(1):e220–e220. doi:10.5001/omj.2021.91.

[3] Fernández-de-las-Peñas C, Palacios-Ceña D, Gómez-Mayordomo V, Cuadrado ML, Florencio LL. Defining Post-COVID Symptoms (Post-Acute COVID, Long COVID, Persistent Post-COVID): An Integrative Classification. Int J Environ Res Public Health. 2021;18(5):2621. doi:10.3390/ijerph18052621.

[4] Halpin S, O’Connor R, Sivan M. Long COVID and chronic COVID syndromes. J Med Virol. 2021;93(3):1242–1243. doi:10.1002/jmv.26587.

[5] Pezzini A, Padovani A. Lifting the mask on neurological manifestations of COVID-19. Nat Rev Neurol. 2020;16(11):636–644. doi:10.1038/s41582-020-0398-3.

[6] Ferrandi PJ, Alway SE, Mohamed JS. The interaction between SARS-CoV-2 and ACE2 may have consequences for skeletal muscle viral susceptibility and myopathies. J Appl Physiol. 2020;129(4):864–867. doi:10.1152/japplphysiol.00321.2020.

[7] Morgul E, Bener A, Atak M, et al. COVID-19 pandemic and psychological fatigue in Turkey. Int J Soc Psychiatry. 2021;67(2):128–135. doi:10.1177/0020764020941889.

[8] Huang C, Huang L, Wang Y, et al. 6-month consequences of COVID-19 in patients discharged from hospital: a cohort study. Lancet. 2021;397(10270):220–232. doi:10.1016/S0140-6736(20)32656-8.

[9] Xiong Q, Xu M, Li J, et al. Clinical sequelae of COVID-19 survivors in Wuhan, China: a single-centre longitudinal study. Clin Microbiol Infect. 2021;27(1):89–95. doi:10.1016/j.cmi.2020.09.023.

[10] Blomberg B, Mohn KG-I, Brokstad KA, et al. Long COVID in a prospective cohort of home-isolated patients. Nat Med. June 2021. doi:10.1038/s41591-021-01433-3.

[11] Davis HE, Assaf GS, McCorkell L, et al. Characterizing Long COVID in an International Cohort: 7 Months of Symptoms and Their Impact. medRxiv. January 2021:2020.12.24.20248802. doi:10.1101/2020.12.24.20248802

[12] Mandal S, Barnett J, Brill SE, et al. ‘Long-COVID’: a cross-sectional study of persisting symptoms, biomarker and imaging abnormalities following hospitalisation for COVID-19. Thorax. 2021;76(4):396–398. doi:10.1136/thoraxjnl-2020-215818.

[13] CarfÌ A, Bernabei R, Landi F. Persistent Symptoms in Patients After Acute COVID-19. JAMA. 2020;324(6):603. doi:10.1001/jama.2020.12603.

[14] Sykes DL, Holdsworth L, Jawad N, Gunasekera P, Morice AH, Crooks MG. Post-COVID-19 Symptom Burden: What is Long-COVID and How Should We Manage It? Lung. 2021;199(2):113–119. doi:10.1007/s00408-021-00423-z.

[15] Hopkins C, Surda P, Whitehead E, Kumar BN. Early recovery following new onset anosmia during the COVID-19 pandemic – an observational cohort study. J Otolaryngol - Head Neck Surg. 2020;49(1):26. doi:10.1186/s40463-020-00423-8.

[16] D’Ascanio L, Pandolfini M, Cingolani C, et al. A. Olfactory Dysfunction in COVID-19 Patients: Prevalence and Prognosis for Recovering Sense of Smell. Otolaryngol Head Neck Surg. 2021 Jan;164(1):82–86. Doi: 10.1177/0194599820943530.

[17] Stefano GB, Büttiker P, Weissenberger S, Martin A, Ptacek R, Kream RM.Editorial: The Pathogenesis of Long-Term Neuropsychiatric COVID-19 and the Role of Microglia, Mitochondria, and Persistent Neuroinflammation: A Hypothesis. Med Sci Monit. 2021 May 10;27:e933015. Doi: 10.12659/MSM.933015.

[18] Pinna P, Grewal P, Hall JP, et al. Neurological manifestations and COVID-19: Experiences from a tertiary care center at the Frontline. J Neurol Sci. 2020;415:116969. doi:10.1016/j.jns.2020.116969.

[19] Karadaş Ö, Öztürk B, Sonkaya AR. A prospective clinical study of detailed neurological manifestations in patients with COVID-19. Neurol Sci. 2020;41(8):1991–1995. doi:10.1007/s10072-020-04547-7.

[20] Romero-Sánchez CM, Díaz-Maroto I, Fernández-Díaz E, et al. Neurologic manifestations in hospitalized patients with COVID-19: The ALBACOVID registry. Neurology. 2020;95(8):e1060–e1070. doi:10.1212/WNL.0000000000009937.

