## Supplementary material for "Long COVID in hospitalized and non-hospitalized patients in a large cohort in Northwest Spain, a prospective cohort study": Questionnaire

**SUPPLEMENTARY DATA**

**Questionnaire of follow-up after COVID-19 disease**

- **Blood type**:

A+	A-	B+	B-	AB+	AB-	0+	0-

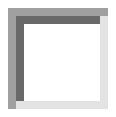

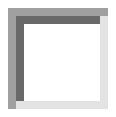

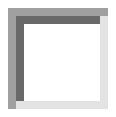

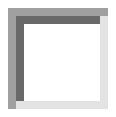

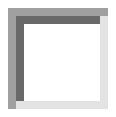

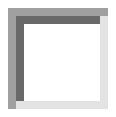

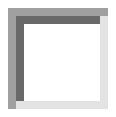

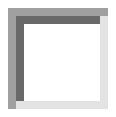

- **Evaluation corresponding to**:

1st Month	3rd Month	6th Month	12th Month	Another date:

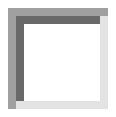

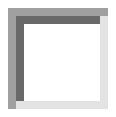

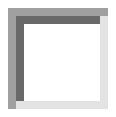

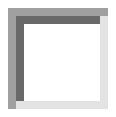

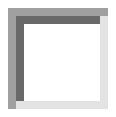

- **Persistence of symptoms**:
  - No

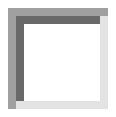

- Yes

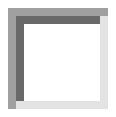

- Type of symptoms:
  - **General**

Fatigue	Sore throat	Hair loss	Insomnia

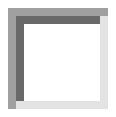

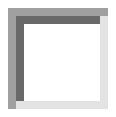

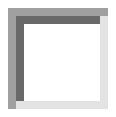

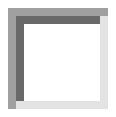

- **Neurologic**:

Loss of smell	Loss of taste	Headache	Tingling	Numbness	Dizziness	Confusion

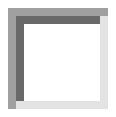

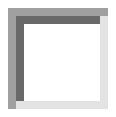

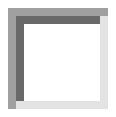

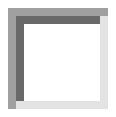

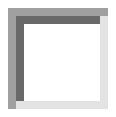

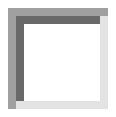

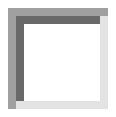

- **Eye**:

Conjunctivitis	Dry eye	Chemosis	Epiphora

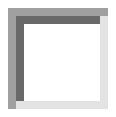

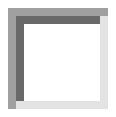

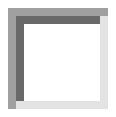

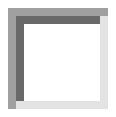

- **Thoracic**:

Cough	Chest pain

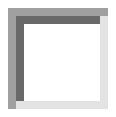

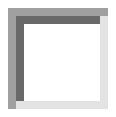

Dyspnoea	→ Grade (mMRC Scale): 1	2	3	4

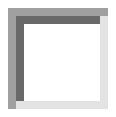

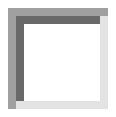

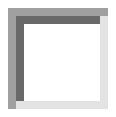

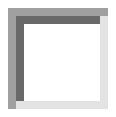

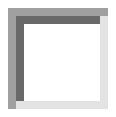

- **Digestive**:

Diarrhoea	Nausea	Constipation	Abdominal pain

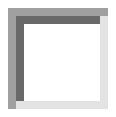

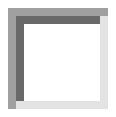

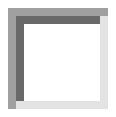

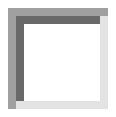

- **Musculoskeletal**:

Muscle pain	Joint pain

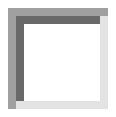

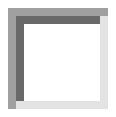
